# Exploratory Genome Sequence Analysis of Candidate Genes Identified Three Loci Potentially Related to Mefloquine Side Effects

**DOI:** 10.64898/2026.07.11.26357837

**Authors:** Monique Hollis-Perry, Jeffrey Livezey, Daoqin Bi, Joshua Gray, Dutchabong Shaw, Daniel Hupalo, Milissa U. Jones, Heidi Adams, Priscilla Kobi, Xijun Zhang, Karl C. Alcover, Lydia D. Hellwig, Louis R. Cantilena, Matthew D. Wilkerson, Clifton Dalgard, David Saunders

## Abstract

**BACKGROUND:** Despite effectiveness as a once-weekly antimalarial prophylaxis, mefloquine has fallen out of favor due to its neuropsychiatric side effects. While possible genetic susceptibilities have been identified in preliminary studies, pharmacogenomic testing guidance is not available for mefloquine.

**METHODS:** Volunteers with a history of mefloquine exposure were recruited to a cross-sectional case-control study. Pharmacogenomic analysis was performed on 7 candidate genes of interest with 16 missense variants including *ORM1, MTHFR, MDR1, PYK2, HT2A, ADA, and ADORA2A*.

**RESULTS:** Fifty participants enrolled including those who had mefloquine exposure and chronic adverse effects (AEs) lasting 6 months or longer (n = 23); with subsequent AEs less than 6 months (n = 12); no AEs (n = 8); and a control group with a history of post-traumatic stress disorder (PTSD) but no mefloquine exposure (n = 7). Psychometric testing showed that mefloquine users with AEs lasting 6 months or more and PTSD patients who had not used mefloquine reported more evidence of sleep impairment, balance and equilibrium disorders, and lower levels of psychological well-being than mefloquine users who reported without AEs or with AEs but lasted less than 6 months. The *ADORA2A* gene was found to carry a higher burden of variation among volunteers exposed to mefloquine with AEs compared to those who did not. The variant rs141942830 within *ADORA2A* was observed to be higher among cases compared to the reference allele frequency listed in the gnomAD database but was found to not be significantly enriched. In addition, *MTFHR* gene was found to be enriched for variation in volunteers with long-term side effects compared to those with short-term or no side effects.

**CONCLUSIONS:** Volunteers who reported long-term adverse events after exposure to mefloquine had excess rare variation within the *ADORA2A* gene compared to those without adverse events and those with short term adverse events. The *ADORA2A* rs141942830 was identified as a new variant of interest, as it was elevated but not significantly enriched among cases of long-term AEs, compared to the population frequency reported by gnomAD. These non-silent variants may serve as mediators to alternate pathways for signal transduction or drug metabolism.

## BACKGROUND

Mefloquine, a 4-quinolinemethanol synthetic quinoline, was approved by U.S. Food and Drug Administration (FDA) in 1989 for prevention and treatment of malaria. Due to risk of serious neuropsychiatric adveres reactions reported, FDA added a “black box” warning to the label in 2013 (1). Direct evidence published in 2006 found that mefloquine is neurotoxic, causing brain stem lesions that are “permanent in nature” in animal models (2). Currently mefloquine is recommended as an alternative antimalaria drug for use with caution. Nevertheless, it continues to be used frequently as an alternative against *P. falciparum* malaria with multi-drug resistance areas. However, malaria prevention efforts remain challenged by not only population migration and climate change but also emerging resistance to antimalarial drugs (4,5). Ensuring compliance with medication use, minimizing logistical barriers, and overcoming patient reluctance to disclose mental health concerns also challenge efforts to ensure safe and effective malaria prevention and therapy (6–9).

Neuropsychiatric symptoms associated with mefloquine use include dizziness, headaches, memory impairment, anxiety or depression, hallucinations or vivid dreams or nightmares, and psychosis (10–16). Clinical risk factors for complications include female sex; first-time users, low body mass index, and personal or family history of seizure or psychiatric disorder (17,18). Symptoms may vary in severity, develop after the first or second dose, and do not correlate with either gender or levels of drug or metabolite in serum or plasma (19–21). In cases with symptoms of nausea, dizziness, swaying, and unsteadiness, the differential diagnosis would include other causes of vestibular disorders (2,22,23).

Individuals with neuropsychiatric adverse events (AEs) due to mefloquine use may have risk factors that, if pre-emptively identified, can provide a signal to select an alternative malaria prophylaxis regimen. Studies in military service members deployed to Iraq and Afghanistan illustrated how extrinsic circumstances, particularly combat experiences and compliance with safety guidelines, can introduce confounding factors to a rigorous assessment of a medication’s utility in selected populations (24–26). The overlap between symptoms seen in mefloquine toxicity include post-traumatic stress disorder (PTSD) and disruptions in executive and visuospatial functioning has been noted by other investigators (27–30).

To assess the potential association between antimalarial drug exposure and health outcomes, an ad hoc committee was formed by the National Academies of Sciences, Engineering, and Medicine. Through reviewing available literature, reports, and documents and testimony from experts and affected individuals and advocates, the committee concluded that the potential long-term health effects remain unclear, and more research is warranted (31). The aim of the present study was to seek a better understanding of the potential link between mefloquine exposure and neuropsychiatric symptoms among at-risk service members. Genotyping was used to correlate cellular targets and neurotransmitter pathways associated with clinical symptoms of mefloquine toxicity to identify potential targets for clinical test development. This case-control, cross-sectional study analyzed possible associations between genomic variants associated with symptoms and neuropsychological testing results to determine whether pharmacogenomic guidelines can be developed to more precisely guide future mefloquine use (32,33). Volunteers with a history of exposure to mefloquine, with or without adverse neuropsychiatric symptoms, were invited to participate based on medical review of past mefloquine exposures. A control group of volunteers who were not exposed to mefloquine but had been diagnosed with PTSD without prior traumatic brain injury (TBI) was included.

## METHODS

### Study Design

Investigators combined a clinical interview, self-administered neuropsychiatric tests, and analysis of selected genetic polymorphisms to assess differences between groups. A total of 53 volunteers between 24 and 74 years of age were enrolled. The study was approved by the Uniformed Services University of the Health Sciences Institutional Review Board under protocol MED-83-9137. All participants or their legally authorized representatives provided informed consent to participate in the study. Psychometric testing and blood sample collection were completed between October 24, 2019 and July 19, 2022.

### Volunteer Enrollment

Eligibility criteria for prospective volunteers included: travel to an area with malaria prophylaxis recommendation at time of prophylaxis; and a history of weekly malaria prophylaxis with at least one dose of mefloquine. Volunteers with symptoms of mefloquine neurotoxicity were divided into two groups based on duration of symptoms after stopping mefloquine use. Volunteers in Groups 1 and 2 (G1 and G2) reported symptoms lasting six months or longer and less than six months, respectively. Two negative control groups consisted of volunteers who met criteria for mefloquine exposure but denied any neuropsychiatric symptoms (Group 3, G3), and volunteers who were diagnosed with PTSD but had no history of mefloquine exposure or traumatic brain injury (Group 4, G4). Symptom duration was estimated by start and end dates reported by volunteers. If volunteers reported that symptoms persisted, the date of study enrollment was selected to estimate symptom duration.

Volunteer demographic and interview information was transcribed from paper case report forms into the REDCAP data collection system. Symptom duration was calculated in Excel (Version 2312 Build 16.0.17126.20132).

### Study Procedures

#### Psychometric Tests

Self-administered computerized tests were selected to provide uniform, reproducible data with test-retest reliability. The PTSD Checklist for DSM-5 items cover common symptoms and are rated on a 5-point Likert scale (34–36). The Patient-Reported Outcomes Measurement Information System (PROMIS) was used to measure and track responses in physical, mental, and social areas, including questionnaires that assess severity and frequency of sleep impairments (37,38). To capture symptoms related to disequilibrium, we selected the Dizziness Handicap Inventory Scale (DHI) and Vertigo Symptom Scale—Short Form (VSSF) as standardized and validated methods (39,40). The NIH Toolbox Cognition Battery was used to survey attention, episodic and working memory, language, executive function, and processing speed (41). In this study, selected elements of the NIH Toolbox Emotion Battery that assessed elements of anxiety including affect, psychosocial functioning, peer relations, social perception, and well-being. An overlap between elements in the Emotion Battery and PROMIS tests provides internal validation (42). The Test of Memory Malingering (TOMM), a validated test, was used to identify invalid cognitive test performance (43).

#### Whole Genome Sequencing

Table 1 shows the 7 genes and 16 single nucleotide variants (SNVs) selected for test evaluation. Variants were selected based on prior studies suggesting links to mefloquine toxicity.

Donor’s whole blood samples were collected in PAXGene RNA tubes that were prelabeled with subject identification number and date of collection. The samples were stored in a -80^0^C freezer until analysis. After DNA extraction, samples with a DNA concentration of at least 30 ng/μL were included for further seqencing. Sequencing libraries were prepared from DNA extracted from donors’ whole blood samples which were then sequenced at The American Genome Center at Uniformed Services University. Extracted DNA was quantified, normalized, sheared and prepped using a Spectramax Gemini XS, Covaris LE220, Roche Light Cycler, and Illumina c-Bot2. Whole Genome Sequencing (WGS) was generated using PCR-free tagmentation-based preparation as previously described (44,45). All sequencing runs generated 2 × 150 bp reads and was performed on the NovaSeq 6000 (Illumina) System. Sequencing libraries were multiplexed using unique dual indexes into pools of 24 to 32 libraries for sequencing by using the S4 flow cell with 300 cycle SBS kits.

Data generation was completed on the Illumina HAS Bioinformatics Pipelineincluding processes for alignment, variant calling for copy number and structural variants, and annotation using the GRCh38 human reference genome (45). Ancestry and biological sex was measured using Peddy software and principal component analysis (PCA) compared to self-reported values (46,47). Whole-genome variation calls from each sample was used for subsequent statistical analysis after limiting to the variants of interest.

#### Analysis of Genetic Determinates of Mefloquine Sensitivity

A Fischer’s Exact test of allele counts was used to compare the frequency of different alleles at loci of interest between cases and controls (i.e., associations with mefloquine response or related symptoms compared to individuals with no adverse events). In addition, a logistic regression test looked for an increased frequency of certain alleles among the mefloquine cases compared to the controls, adjusting for biological sex and ancestry. To consider the total burden of the variants within each gene of interest we tested using a gene-collapsed SKAT-O paradigm, as well as testing single-variants for association with the mefloquine sensitivity phenotype, as implemented in the software EPACTS (https://github.com/statgen/EPACTS). The first four eigen vectors of the PCA and the genetically inferred biological sex were used as covariates for all single variant and SKAT-O analyses. All samples collected from subjects with exposure to mefloquine were included in the analysis (N=43). This included comparisons between Group 1 (mefloquine exposure users with symptoms lasting 6 months or more) to and Group 3 (mefloquine exposure users with no symptoms) (G1 to G3), and between Groups 1 and 2 (mefloquine exposure users with long- or short-term symptoms) combined with Group 3 (mefloquine exposure users without any symptoms) (G1 + G2 to G3 and G1 to G2 +G3). Tests were limited to the 7 genes of interest and included variants at or below 5% allele frequency.

## RESULTS

### Volunteer Demographics

Most volunteers requiring malaria prophylaxis were visiting countries in Africa. (Figure 1). There was roughly equal representation between males (47.2%) and females (52.8%). The mean age of the volunteers was 39.6 years (+/- 10.4, mean ± SD). The sample mean Body Mass Index (BMI) was 26.1 (+/- 5.1, mean ± SD). Most of the volunteers were non-Hispanic (94.3%) and Caucasian (79.2%) (Table 2). Most volunteers who took mefloquine reported 1 (60.4%) or 2 (18.9%) courses, consisting of 1 tablet weekly. One volunteer took a course of treatment following malaria infection. Educational attainment ranged from high school (2; 3.8%), some college or associate’s degree (3; 5.7%), bachelor’s degree (12; 22.6%), to advanced degree (36; 67.9%). Rates of both reported alcohol use and exposure to traumatic events while taking mefloquine were highest in Group 2 (86.7% and 53.3%, respectively). No volunteers in Groups 1-3 had medical or behavioral conditions that would have precluded mefloquine prophylaxis prior to use. Overall rates of reported alcohol use and traumatic exposures were higher for females in Group 2 than Group 1. Males in Group 2 had a higher reported rate of alcohol use than those in Group 1. The reporting rates for traumatic events among the groups were similar.

### Symptom Types and Frequency

Clinical symptom categories reported by volunteers included disorders related to sleep, anxiety, emotion and mood, trauma and stress response, suicide-related behavior, attention and memory, equilibrium, and perception (Table 3). Volunteers may report more than one symptom within a category or report a symptom occurring during more than one exposure to mefloquine (Groups 1 and 2) or trauma (Group 4). One volunteer reported vivid dreams during each of three exposures. The three most frequently reported symptom types were related to sleep (40 reports); emotion and mood (22 reports); and anxiety, panic attacks and phobias respectively (20 reports, Table 3). In both groups, volunteers may have reported more than one symptom or one symptom during more than one exposure. Symptoms related to disturbances of emotion, mood, or affect; sleep; attention or memory; dizziness or equilibrium; neuropathy; motor function were also reported. Symptoms lasted up to years after mefloquine use ended and, for several cases, remained current when the volunteers were interviewed. Symptoms reported by 2 or more volunteers in Group 2 were also reported by those in Group 1, but in lower numbers and percentages of overall mefloquine exposures.

Group 1 volunteers (9 males; 14 females) reported symptom onset within 3 months of starting and lasting at least 6 months after ending a course of mefloquine for malaria prophylaxis or therapy. Group 2 volunteers (6 males; 9 females) reported symptoms lasting from within 3 months of starting to fewer than 6 months after finishing mefloquine prophylaxis or therapy. Group 3 volunteers (4 males; 4 females) reported no symptoms occurring within 3 months of starting a course of mefloquine. Group 4 volunteers (6 males; 1 female) reported symptoms consistent with their prior diagnoses of PTSD and resembling those seen in groups 1 and 2 (e.g., nightmares, hypervigilance). They did not report symptoms linked to vestibular system disorders (e.g., tinnitus, vertigo), speech and language, neuropathic or neuromuscular symptoms, or specifically related to pain. Group 4 volunteers share some, but not all, symptoms reported by Groups 1 and 2: disorders in sleep, attention and memory, emotion, or mood were reported; but those related to balance and equilibrium were not (Table 3).

### Genomic Analysis

Blood samples from 50 volunteers were collected and completed whole-genome sequencing. Ancestry of included individuals ranged from European, Native American, African, East Asian, and South Asian. Biological sex, determined genetically for each sample, showed approximately equal male-female distribution (25 males; 28 females) (Table 2). Group combinations for testing compared presence or absence of symptoms; symptom chronicity; and long-term symptoms reported by those who used mefloquine versus those who did not. Specifically, investigators compared groups reporting long-term symptoms (G1) with those reporting no symptoms (G3) as the primary test (G1 to G3). Secondarily, volunteers who used mefloquine and reported long- or short-term symptoms were compared with those reporting none: (G1 and G2) to G3. A third group, mefloquine users with long-term symptoms were compared to mefloquine users reporting either short-term or no symptoms and those who had not used mefloquine: G1 to (G2 and G3). Lastly, mefloquine users with long-term symptoms (G1), was compared to unexposed individuals with PTSD (G4) (G1 to G4).

Given the limited sample size for testing, our analysis was exploratory in nature. Using a variety of testing strategies, we aimed to identify any enrichment of genetic variation within a set of known genes of interest. Using a Fisher’s Exact test to perform the comparisons described above, we have not found any significant difference between any comparison of groups. Similarly, after performing logistic regression for the groups described above, we found no significant difference between any comparison of groups.

Using the SKAT-O test to compare individuals with long-term symptoms to those with short-term or no symptoms, we found that the gene of interest *ADORA2A* was significant for an enrichment of rare variation after adjusting for Bonferroni multiple testing in both comparisons (p < 0.0007) (Table 4). No single variants were found to be significantly enriched within this gene, however the allele frequency of the *ADORA2A* variant rs141942830 (p. Val275Ile) was observed to be higher in long term symptomatic cases than the allele frequencies reported in the gnomAD database (rs141942830: 0.021739 group 1 cases / 0.000317 gnomAD).

In addition, the gene of interest *MTHFR* was identified as significantly enriched for a rare variation by SKAT-O when comparing G1 to (G2 + G3). However, this result was not replicated in other group comparisons. No notable single variants were observed within *MTHFR*, however in the Fisher’s Exact test, we identified one near-significant variant. *MTHFR* variant rs1801131 approached significance (p=0.0875 and 0.064) when comparing long-term symptoms to no symptoms after mefloquine exposure (G1 to G3 p=0.0875) and long-term symptoms compared to all other individuals exposed to mefloquine (G1 to G2 and G3, P=0.064), respectively. When comparing the allele frequency of long-term mefloquine symptoms (0.3043) to the population frequencies present in the gnomAD database for the variant rs1801131 (0.2634), we found that this variant was not significantly enriched in the long-term symptoms group (G1) compared to the global population.

### Psychometric Test Results

Test results of 52 volunteers validated by TOMM assessment (passed rate of 98%) were included in subsequent analysis. Volunteer who had not used mefloquine (Group 4) had higher median PROMIS scores for sleep-related impairment. Among mefloquine users, those with long-term symptoms had the highest median scores for sleep disturbance and sleep-related impairment (Table 5). As expected, the DSM-PTSD Checklist Total median score for the control group with PTSD (Group 4) was higher than those of groups that had used mefloquine. Additionally, this control group’s median Psychological Well-Being Summary score was lower than those of the other groups. Among mefloquine users, DSM-PTSD median scores were higher in Group 1 than in Groups 2 and 3. Psychological Well-Being Summary scores were lower in Group 1 than in Groups 2 and 3. Mefloquine users in Group 1 reported higher median values on the DSM-5 PTSD Checklist and Total DHI than those in Groups 2 or 3. P-values were < 0.0001 and 0.0055, respectively using one-way analysis of variance (ANOVA) (Figure 2) (48).

Additionally, the Vertigo Symptom Scale—Short Form (VSSF) median score for volunteers in Group 1 was higher than those reported for both Groups 2 and 3. Preliminary analysis of PROMIS data for anxiety and anger showed higher median scores in Groups 1, 2, and 4 (volunteer groups reporting symptoms related to mefloquine and PTSD controls) compared to those who reported no symptoms after mefloquine use (Group 3). Cognition Total Composite Score median values were higher in Groups 2 and 3 than in Groups 1 and 4.

Volunteers’ responses to individual test questions were compared with genotype data (Supplementary Tables). Five single nucleotide polymorphisms (SNPs) were found to be associated with the Dizziness Handicap Inventory Score. Specifically, one question in the emotional domain (“Because of your problem, are you depressed?”) was associated with *MDR1* rs1045642 and HT2A rs7997012. Most volunteers with heterozygous genotypes responded “No”. Additionally, two physical domain questions addressing difficulty walking due to symptoms were found to be associated with *MTHFR* rs1801133 and *ORM1* rs1126801. Most volunteers with homozygous reference or heterozygous genotypes answered “No” to both questions. Moreover, associations between SNPs of interest and two functional domains questions were identified. One of which questions addressing movement disturbance at home was associated with ORM1 rs17650, the second question assessed travel restrictions was associated with *MTHFR* rs1801133. Most respondents who reported no travel restrictions had a homozygous reference genotype.

## DISCUSSION

Potential pharmacogenomic targets previously linked to symptoms reported by patients after mefloquine use were selected to determine whether an association with mefloquine-related side effects could be identified. The study yielded new insights on possible genetic associations with mefloquine toxicity. Our exploratory genomic analysis revealed that volunteers with long-term AEs after taking mefloquine had excess rare variation within the *ADORA2A* gene compared to those without AEs, and those with short term AEs. We also identified rs141942830 as a new variant of interest, as it was elevated among cases with long-term AEs, compared to the population frequency reported by the gnomAD database. These non-silent variants may serve as mediators to alternate pathways for signal transduction or drug metabolism.

Clinical findings from the current study were consistent with previous reports. Female volunteers or those with lower BMI were the majority of those who reported neuropsychiatric symptoms lasting six months or longer (Group 1) (21). As reported in prior studies symptoms related to vestibular disorders or associated conditions (e.g., dizziness, problems with balance, tinnitus) were observed in Groups 1 and 2, which could be resulted from MQ-related ototoxicity (16,49,50).

### Genetic Variants Evaluated in This Study

#### Alpha_1_-acid glycoprotein (*AAG*/ORM1)

MQ distribution in the brain is determined by quantification of the alpha_1_-acid glycoprotein (AAG), primarily the orosomucoid 1 (ORM1) variant (51). AAG is an acute-phase reactant that binds receptors on multiple cell types, transports ligands in response to inflammation, and can be elevated in cancer and sepsis (52,53). Immune-modulating functions of the AAG occur through interaction with cytokines and include macrophage activation(54). Studies with propranolol and lidocaine demonstrated the association between AAG and blood brain barrier (BBB) transit (55,56). Researchers have observed ORM1 polymorphism expression variations among ethnic groups and interindividual responses to medications of different drug classes (57–62). The *ORM1*S*, *ORM1*F1*, and *ORM1*F2* alleles (rs17650, rs1126801) share A to G transitions in position 20 in exon 1 and position 156 in 5 of the *AAG* gene on chromosome 9 (63). Health status and intervening clinical factors (e.g., age, inflammatory states, pregnancy, and cancer) impact *AAG* levels (64–66).

#### ATP-dependent p-glycoprotein (P-gp/*MDR1*)

The ATP-dependent p-glycoprotein (P-gp) efflux pump transports several different drug types across cellular membranes including the BBB, intestine, and liver. It is encoded by *MDR1*, (multidrug resistance gene), also known as the human *ABCB1* gene (ATP-binding cassette, subfamily B), which is located on chromosome 7q21.1 (67–69). In the central nervous system (CNS), P-gp is expressed primarily on the BBB luminal membranes of capillary endothelial cells but has also been localized to astrocytes and microglia (70–72). The P-gp receptor interacts with multiple cellular processes, and its number or function may be altered by infection, inflammation, or malignancy (73,74). P-gp expression on circulating and mucosa-associated cells contributes to immune surveillance and may impact disease development and progression(75). P-gp efflux drug movement from the cytoplasm to the extracellular space can lead to a sub-therapeutic response or a treatment-resistant strain of *Plasmodium falciparum* (76,77). MQ can inhibit this multidrug pump by blocking efflux at the intestine and BBB (15,78–81). A year-long prospective cohort study documented volunteers reported adverse events (AEs), changes in hand-eye coordination by a validated neuropsychiatric test, and trends of elevation in serum levels of MQ and its primary metabolite. It was found that 3 variants (C1236T, G2677T, and C3435T) and the 1236-2677-3435 TTT haplotype were associated with neuropsychiatric AEs, particularly in female volunteers (82).

#### Pyk non-receptor tyrosine kinase (*Pyk2*/*PTK2B*)

Proline-rich tyrosine kinase 2 (*Pyk2*) is one of two members of the focal adhesion kinase (*FAK*) family cytogenetically located on 8p21.2 (83,84). Other names of this non-receptor tyrosine kinase include *PTK2B*, *RAFTK* (related adhesion focal tyrosine kinase), *CATK* (calcium-dependent tyrosine kinase), *CAKβ* (cell adhesion kinase β), and *AKT* kinase (85–87). *Pyk2* functions include roles in cell signaling and stress response processes, including mitogen-activated protein (MAP) kinase activation (88–92). *Pyk2* induction may regulate angiogenesis, vascular integrity, and platelet adhesion through endothelium-derived nitric oxide (eNOS) activation (93,94). In the CNS *Pyk2* is preferentially expressed in the hippocampus, cerebral cortex, and olfactory bulb. N-methyl D-aspartate (NMDA) receptor phosphorylation regulates anti-apoptotic mechanisms, hippocampal neural migration, learning, and memory development (86,95–97). Ischemia, inflammation, and other environmental shifts alter *Pyk2* function and induce endothelial cell changes. Overexpression of *Pyk2* can lead to apoptosis and promote tumor development and progression (98–100). Genome-wide association studies (GWAS) identified rs28834970 as a susceptibility locus for Alzheimer’s disease (AD) (101,102). MQ neurotoxicity occurring through *Pyk2*-related pathways results from similar disruptions in calcium movement, oxidative stress, and apoptosis (103–106).

#### Methylenetetrahydrofolate Reductase (*MTHFR*) Enzyme

*MTHFR* regulates cellular homeostasis by converting homocysteine to methionine and providing methyl groups to DNA modification (107,108). *MTHFR* activity prevents homocysteine accumulation, which has been linked to oxidative stress, DNA damage, and apoptosis initiation (109–112). Homocysteinemia, a group of hereditary disorders due to one of several *MTHFR* enzyme deficiencies, is associated with cardiovascular disorders with variable severity and age of onset (113–115). Two gene variants, located on chromosome 1p36.22, include C677T (rs1801133; alanine to valine), and A1298C (rs1801131; glutamate to alanine) (116). Reported causes of impaired *MTHFR* function in C677T homozygotes include impaired DNA methylation due to suppressed gene expression and lower enzyme levels (117–121). C677T distribution varies by ethnicity and geography . Proposed associations between *MTHFR* polymorphisms cardiovascular symptoms, thrombotic disease, and neuropsychiatric disorders lack clear consensus in the literature, which may reflect effects of conflicting evidence, overlapping cellular pathways, environmental or other confounding factors (122–126).

Although no clinical studies explicitly connect *MTHFR* variants with mefloquine neurotoxicity, TBI, or PTSD, they may share the same or similar metabolic pathways (108,127–130). Homocysteine, a proposed intermediary, can indirectly regulate levels of neurotransmitters, overstimulate NDMA receptors, or increase oxidative stress through lipid peroxidation, electrolyte disruptions, and neuronal death (131–134).

#### 5HT2A (Serotonin) Receptor

Serotonin (5-hydroxytryptamine, 5HT) is a neurotransmitter that regulates diverse physiologic and behavioral functions by interacting with a family of receptors characterized by structure and pharmacologic traits (135–137). Receptor functions include dopamine and NDMA receptor agonist mechanisms that are implicated in schizophrenia (138–141). The *5HTR2A* gene is mapped to chromosome 13q14-21 (142). Variants evaluated in this study include: rs7997012, rs1928040, rs6311, and rs6313. An association between SNPs rs6311 and rs6313 and development of schizophrenia has been reported (143,144). A recent study reported that *HTR2A* SNPs rs7997012 and rs6311 could predict response to atypical antipsychotics (145). Comparisons of genotypes and treatment response identified an association in patients with genotype rs7997012 and a more favorable treatment outcome after antidepressant therapy. In groups with the genotypes rs1928040, rs6311, and rs6313 evidence of an association with antidepressant treatment is less robust (146,147). Elements of a mechanism to explain potential therapeutic applications of psychotomimetic drugs have been observed in mefloquine users. Lipophilicity of MQ facilitate it’s access into neurons and affinity with *5HT2A* receptors (73,148–150).

#### Adenosine deaminase (*ADA*)/Adenosine 2-α Receptor (*A2A*)

Adenosine, adenosine deaminase (*ADA*), and its receptors are components of sleep homeostasis. Adenosine counteracts activity in neurons maintaining vigilance and promotes prostaglandin-mediated sleep (151,152). The *ADA* rs73598374 polymorphism (Asp8Asn; G22A) identified in healthy volunteers is associated with reduced adenosine conversion to inosine. Sleep patterns of A allele carriers showed deeper and more efficient sleep cycles (153–156). Caffeine, an adenosine deaminase receptor antagonist used as a stimulant to improve alertness or mitigate effects of sleep deprivation, also has anxiogenic effects which may be moderated by *ADORA2A* genotypes (157–159).

The *ADORA2A* receptor (*A2A*) polymorphisms rs5751876 (T1976C; 1083C/T) and rs35320474 (C2952T) affect sleep quality and duration (160). Subjective insomnia was reported more frequently in caffeine-sensitive volunteers who were more likely to be C/C homozygotes. T allele carriers may be insensitive some anti-A2AR effects(160–162). Rs5751876 (T1976C; 1083C/T) is associated with development of anxiety and related disorders, symptom manifested from which can be aggravated by caffeine use (163–167). The gene for these SNPs is located on chromosome 22q11.23 (168). Mefloquine is a potent and selective A2A antagonist, which may explain common adverse reactions observed in prior studies (169–171).

### Genomic Analysis

SKAT-O analysis showed that the *ADORA2A* and *MTHFR* genes were enriched for variation within individuals with AEs compared to those who had no adverse events. The variant rs141942830 within ADORA2A was elevated in allele frequency (0.02) compared to the allele frequencies listed in gnomAD (0.0003) but was not found to be significantly enriched. Similarly, rs372335618 was found to be overrepresented in controls but was also not significantly enriched. However, given the exploratory nature of this small sample size we cannot definitively associate these variants with adverse events. Looking more closely at *MTHFR,* it has roles in folate metabolism and DNA methylation (108,172). *MTHFR* polymorphisms have been associated with psychiatric disorders and methotrexate toxicity . Enzymes involved with adenosine metabolism have been linked to anxiety, panic disorder, and sleep cycle variations (154,165,173). Adenosine deaminase receptor polymorphisms also affect responses to caffeine, dopamine, and prostaglandins, which may also result in increased wakefulness or a heightened response to stress (152,174,175). However, the *MDR1*, *5HT2A*, *ORM1*, and *Pyk2* genes and selected variants of interest were not found to have a statistically significant association with mefloquine symptoms reported in this study.

### Psychometric Testing

The significance of observed associations between specific DHIS questions and selected SNPs is unclear. No clear association between other linked or separate genomic or psychometric test results and frequency of symptoms reported is seen (50). Alternatively, a rat model of ototoxic damage illustrates how equilibrium symptoms may develop after mefloquine use (50). This may indirectly explain some of clinical findings (50).

Reported symptoms of nausea, dizziness, and vertigo were loosely correlated with psychometric testing as documented in VSSF and DHI results. Groups 1 and 4 had higher median values than Groups 2 and 3. Problems with sleep measured by PROMIS instruments had similar findings, reflecting symptoms of both mefloquine toxicity and PTSD. During the NIH Toolbox Psychological Well Being Summary volunteers assessed their general life satisfaction, positive motivations for living, and feelings of pleasure and happiness. Higher scores indicate increased levels of well-being (176,177). Groups 2 and 3 had higher median scores than those in Groups 1 and 4 (P=0.0254), suggesting greater levels of well-being reported in volunteers who did not report ongoing symptoms. Concerns whether chronic mefloquine toxicity may confound the diagnosis of PTSD have been reported (19,32). The DSM-5 PTSD Checklist data in our study agrees with these concerns. Groups 1 and 4 had higher median scores than Groups 2 and 3 (P<0.001), which is consistent with reports of adverse health impacts (36). However, statistical significance is lost when comparing Group 1 with Groups 2 and 3 in which those volunteers who also reported a traumatic event were not included (p=0.1419). Although Cognition Total Composite Scores suggest a division between groups experiencing minimal or no symptoms and those with more severe or prolonged symptoms, interquartile ranges have overlapping values, and a p-value > 0.05 does not reach statistical significance.

The demand for access to effective malaria prophylaxis and treatment continues (3,178–182). Effects of population movements and climate change appear in new reports of locally transmitted malaria in non-endemic areas (183,184). As *Plasmodium* species develop resistance to current medications, alternatives may include combinations of current medications or re-introducing previously discontinued regimens. Efforts to control malaria spread have also been hampered by compliance with public health measures. Although successful prophylaxis depends on several factors including Drug choice, dosing frequency, side effects, length and purpose of travel, an optimal regimen is the one most likely to be followed (185–187).

### Study limitations

The study recruitment and enrollment occurred from October 2019 to April 2022 that was affected by logistical constraints in place during the early phase of the COVID-19 pandemic. The current study is subject to limitations related to the low response rate including small sample sizes in all groups of interest. Although volunteers were recruited through both military and civilian networks, demographic distribution across groups was uneven, as seen in Group 4, which has only 1 female volunteer. The small cohort, which has limited generalizability, is not representative of either those individuals who have experienced neurotoxic effects of mefloquine or those who have PTSD. Data analysis is limited by reduced statistical power and might be more easily influenced by outliers. Eliciting information about symptom onset or duration depends on a volunteer’s ability to remember and describe prior events, which may be skewed by recall or self-reporting biases.

## Conclusion

Military operational planning already includes screening measures to identify G6PD-deficient service members for whom primaquine and chloroquine are contraindicated (33,188). Pharmacogenomic testing provides a potential alternative method to identify people at risk for significant neuropsychiatric sequelae not detected during routine medical screening. These may include screening for SNPs associated with PTSD to allow a more rigorous bidirectional assessment of genetic factors that can be used to predict response to mefloquine. However, many proposed biomarkers have not been adequately validated for screening purposes and may need to be used in conjunction with other diagnostic modalities (e.g. screening questionnaires or imaging) to be clinically useful. In addition, to date, the overwhelming majority of these measures have not been proven sufficiently reliable, valid and useful to be adopted clinically (188–190). Future study design may also mitigate inherent pitfalls of retrospective studies, particularly those that investigating potential associations between clinical and laboratory data related to trauma (191–193). This study can serve as a guide to future prospective clinical trials targeting or combining psychometric and pharmacogenomic tools to develop and validate a model based on social and biophysical traits seen in trauma and similar neuropsychiatric conditions to identify risk factors for developing complications of mefloquine therapy.

## Supporting information

Tables and Charts

## Data Availability

All data produced in the present study are available upon reasonable request to the authors

