## Supplementary material for "Exploratory Genome Sequence Analysis of Candidate Genes Identified Three Loci Potentially Related to Mefloquine Side Effects": Tables and Charts

**Table 1 Candidate Genes Selected for Pharmacogenomic Analysis**

| **Gene** | **Variant** | **RSID** | **Chromosome** | **Description** |
| --- | --- | --- | --- | --- |
| Alpha_1_-Acid Glycoprotein (AAG) (Human orosomucoid) | | | 9 | Blood-brain barrier permeability. The association between AAG binding and crossing the BBB was shown with lidocaine and propranolol, 2 lipophilic drugs highly bound by ORM1. Subjects with ORM1 S/S variant are postulated to have higher levels of bound MQ, a decrease in volume of distribution, slower apparent clearance due to alterations in intrinsic clearance, and potentially higher CNS levels. |
| ORM1 | S |  |  |  |
| ORM1 | F1 | rs17650 |  |  |
| ORM1 | F2 | rs1126801 |  |  |
| MTHFR Enzyme | | | 1 | Folate metabolism, DNA methylation, reduced oxidative stress. A link between MTHFR mutation and psychological disorders (e.g., depression) has been suggested, despite conflicting evidence in the literature. No studies link MQ or PTSD and MTHFR mutations. One case of chronic MQ toxicity associated with C677T MTHFR mutation. |
| MTHFR | A1298C | rs1801131 |  |  |
| MTHFR | C677T | rs1801133 |  |  |
| MDR1 | | | 7 | P-glycoprotein ATP-dependent efflux pump. MQ is a substrate and inhibitor of the p-glycoprotein efflux pump MDR-1. MDR-1 polymorphisms may contribute to neuropsychiatric side effects reported predominantly in women. |
| MDR1 | C1236T | rs1128503 |  |  |
| MDR1 | G2677T | rs2032582 |  |  |
| MDR1 | C3435T | rs1045642 |  |  |
| Pyk2 | | | 8 | Non-receptor tyrosine kinase 2; possible MQ target in the brain; hippocampus; association with memory formation and Alzheimer’s. Pyk2 stress; activates endothelial Nitric Oxide Synthase (eNOS), AKT kinase, mobilizes intracellular Ca^2+^. Rat neuronal suppression—MQ-induced cell death. |
| Pyk2 | rs2883490 | rs28834970 |  |  |
| 5HT2a Receptor | | | 13 | Serotonin 5HT2a receptor; associated with psychosis and mood disorders. MQ has a strong affinity for the 5HT2a receptor; psychosis from MQ use may be linked. 5HT2a receptor antagonism is a component of atypical antipsychotic treatment for schizophrenia and plays a role in depression and mood disorders. MQ is a 5HT2a agonist, like many hallucinogens and psychotomimetics like LSD. |
| 5HT2A | rs7997012 | rs7997012 |  |  |
| 5HT2A | rs1928040 | rs1928040 |  |  |
| 5HT2A | rs6311 | rs6311 |  |  |
| 5HT2A | rs6313 | rs6313 |  |  |
| Adenosine 2-α Receptor (A2A)/  Adenosine deaminase (ADA) | | | 20/22 | Adenosine deaminase; associated with anxiety and insomnia. Associations with polymorphisms for sleep and anxiety, two of the most common adverse events attributed to MQ. Limited research to date on links to MQ toxicity. |
| ADA | G22A | rs73598374 |  |  |
| ADORA2A | T1976C | rs5751876 |  |  |
| ADORA2A | C2592T | rs35320474 |  |  |

(References provided in text)

**Table 2 Volunteers’ Demographic Summary**

| **Demographic Characteristic** | Group 1 mefloquine exposure, ongoing symptoms ≥ 6 months | Group 2 mefloquine exposure, ongoing symptoms < 6 months | Group 3 mefloquine exposure, no symptoms | Group 4  Post-traumatic stress disorder without mefloquine exposure | Totals (%) |
| --- | --- | --- | --- | --- | --- |
| Age Mean, +/- SD | 41.7 +/- 10.5 | 36.5, +/- 8.0 | 41.7 +/- 10.5 | 34.1 +/- 7.7 | 39.6 +/- 10.4 |
| BMI Mean, +/- SD | 25.8 +/- 5.0 | 27.1 +/- 5.5 | 27.2 +/- 6.3 | 23.9 +/- 3.1 | 26.1 +/- 5.1 |
| Gender # (%)  Male  Female | 9 (39%)  14 (61%) | 6 (40%)  9 (60%) | 4 (50%)  4 (50%) | 6 (86%)  1 (14%) | 25 (47%)  28 (53%) |
| Races Reported  # (%)  African American  Asian  Caucasian  > 1 Reported | 4 (17%)  2 (9%)  16 (70%)  1 (4%) | 2 (13%)  0 (0%)  13 (87%)  0 (0%) | 0 (0%)  0 (0%)  8 (100%)  0 (0%) | 0 (0%)  1 (14%)  5 (71%)  1 (14%) | 6 (11%)  3 (6%)  42 (79%)  2 (4%) |
| Ethnicity  # (%)  Hispanic  Non-Hispanic | 1 (4%)  22 (96%) | 0 (0%)  15 (100%) | 1 (12.5%)  7 (87.5%) | 1 (14%)  6 (86%) | 3 (6%)  50 (94%) |
| Education Level  # (%)  High School Graduate  Total | 1 (4.3%) | 1 (6.7%) | 0 (0%) | 0 (0%) | 2 (3.8%) |
| Some college; no degree  Total | 1 (4.3%) |  |  | 1 (14.3%) | 2 (3.8%) |
| Associate’s degree  Total | 1 (4.3%) | 0 (0%) | 0 (0%) | 0 (0%) | 1 (1.9%) |
| Bachelor's degree  Total | 5 (21.7%) | 4 (26.7%) | 0 (0%) | 3 (42.9%) | 12 (22.6%) |
| Master’s degree  Total | 12 (52.2%) | 9 (60%) | 7 (87.5%) | 2 (28.6%) | 30 (56.6%) |
| Professional (MD, DDS, DVM, LLB, JD)  Total | 2 (8.7%) | 1 (6.7%) | 1 (12.5%) | 0 (0%) | 4 (7.5%) |
| Doctorate degree  Total | 1 (4.3%) | 0 (0%) | 0 (0%) | 1 (14.3%) | 2 (3.8) |
| **# Mefloquine Exposures**  Min, Max  Mean, +/- SD | 1, 2  1.2 +/- 0.4 | 1, 3  1.3 +/- 0.6 | 1, 5  2.4 +/- 1.3 | N/A | 1, 5, 1.4 +/- 0.8 |
| # MQ Exposures (%)  0  1  2  More than 2 | 18 (78.3%)  12 (21.7%)  0 | 12 (80%)  2 (13.8%)  1 (6.7%) | 2 (25%)  3 (37.5%)  3 (37.5%) | 7 (100%) | 7 (100%) |
| # Mefloquine Exposures  Min, Max  Mean, +/- SD | 1, 2  1.2 +/- 0.4 | 1, 3  1.3 +/- 0.6 | 1, 5  2.4 +/- 1.3 | N/A |  |
| # MQ Exposures Reported by Volunteers (%)  0  1  2  More than 2 | 18 (78.3%)  12 (21.7%)  0 | 12 (80%)  2 (13.8%)  1 (6.7%) | 2 (25%)  3 (37.5%)  3 (37.5%) | 7 (100%) | 7 (100%) |
| Total Duration MQ Exp Days  Min, Max  Mean, +/- SD | 13, 1216  367.0 +/- 388.3 | 30, 915  363.5 +/- 280.4 | 121, 5606  1157.9 +/-1822.8 | N/A | 15, 5606, 503.4 +/- 840.9 |
| # Volunteers reporting Trauma Exposure on Mefloquine | Group 1 | Group 2 | Group 3 | Group 4 | Totals (%) |
| Yes (%)  Total | 5 (21.7%) | 9 (60.0%) | 2 (25.0%) | N/A | 16 (34.8%) |
| # Volunteers reporting Alcohol Exposure on Mefloquine | Group 1 | Group 2 | Group 3 | Group 4 | Totals (%) |
| Yes (%)  Total | 12 (52.2%) | 13 (86.7%) | 5 (62.5) | N/A | 30 (65.2%) |

**Figure 1. Volunteers’ Locations Requiring Mefloquine Use for Malaria Prophylaxis or Treatment**

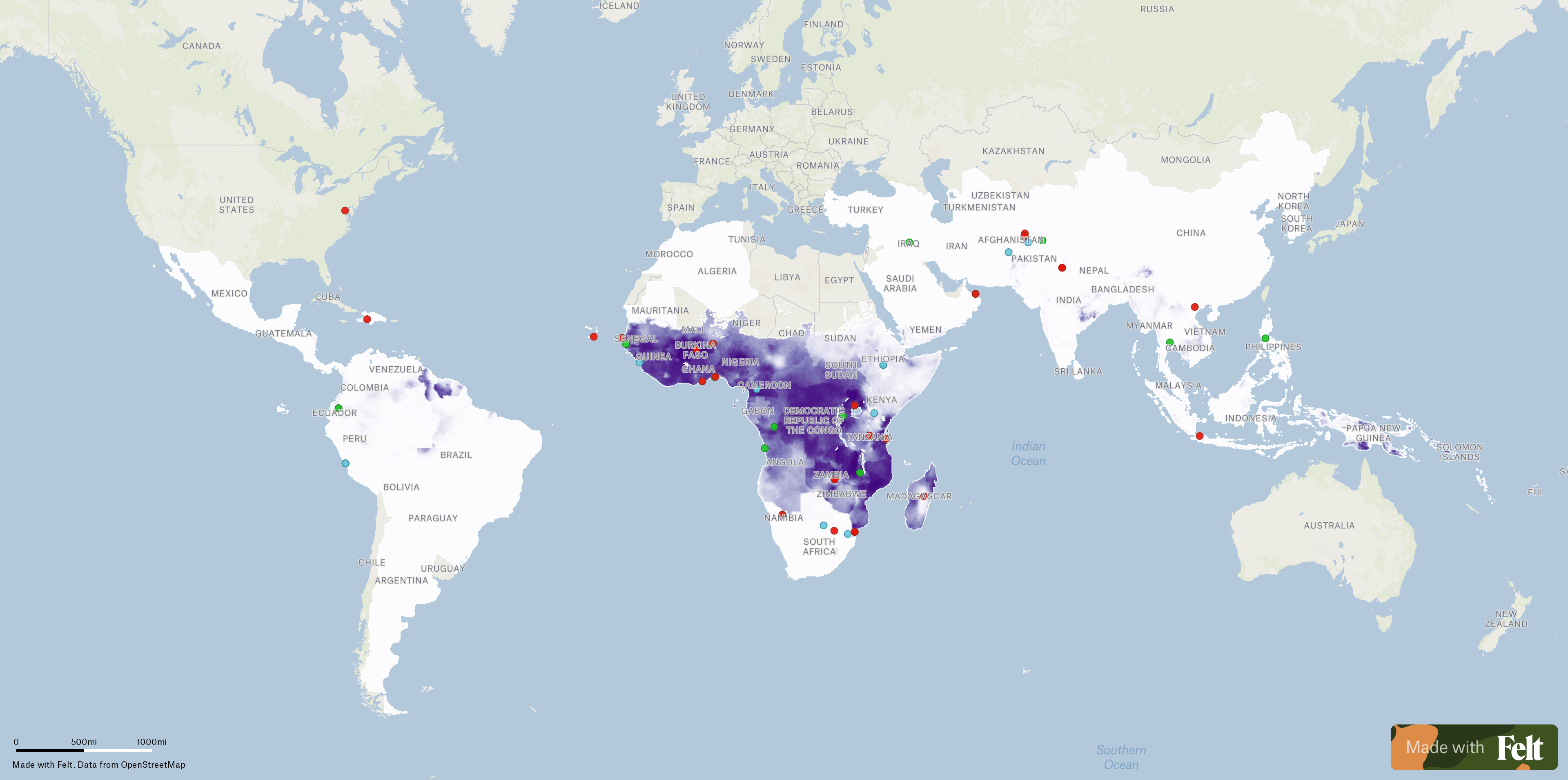

Map generated in Felt

**Table 3: Predominant Symptom Collections**

| **Symptom Type: Total # of Volunteers Reporting Symptoms/Group % of Total** | **Representative Symptoms/Diagnoses** |
| --- | --- |
| Sleep-Related = 40  Group 1: 50.0%  Group 2: 35.0%  Group 4: 15.0% | Vivid dreams, Insomnia, Nightmares, Night Terrors, Parasomnias |
| Emotion, Mood = 22  Group 1: 59.1%  Group 2: 22.7%  Group 4: 18.2% | Aggression, Anger, Depression, Emotional and Mood Lability, Irritability |
| Anxiety, Panic Attacks, and Phobias = 20  Group 1: 70.0%  Group 2: 5.0%  Group 4: 25.0% | Anxiety (unspecified), Panic attacks, Agoraphobia, Claustrophobia |
| Equilibrium = 15  Group 1: 73.3%  Group 2: 26.7%  Group 4: 0.0% | Dizziness, Vertigo, Tinnitus |
| Abnormal Perception = 14*  Group 1: 71.4%  Group 2: 21.4%  Group 4: 7.1% | Hallucinations, Paranoia |
| Attention and Memory = 12  Group 1: 83.3%  Group 2: 0.0%  Group 4: 16.7% | Difficulties with concentration or cognition, Confusion, Memory problems, Impulse control |

*% Totals < 100.0% due to rounding.

**Table 4 Neuropsychiatric Test Score Summary**

| **Survey Instrument** | | **Group 1**  **(Symptoms ≥ 6 months)** | **Group 2**  **(Symptoms < 6 months)** | **Group 3**  **(No Symptoms)** | **Group 4**  **PTSD Only** | **P-value** |
| --- | --- | --- | --- | --- | --- | --- |
| Total Dizziness Score | Median (IQR) | 28  (0-44) | 0  (0-4) | 0  (0, 0) | 4  (0-30) | 0.0055 |
|  | Min-Max | 0-80 | 0-26 | 0-6 | 0-50 |  |
| Cognition Total Composite Score | Median (IQR) | 58.5  (52-63) | 58  (53-63) | 64.5  (60.5-66) | 53  (51-64) | 0.3359 |
|  | Min-Max | 45-82 | 48-74 | 46-72 | 39-78 |  |
| Psychological Well Being Summary | Median (IQR) | 42  (36-47) | 46  (44-59) | 50.5  (45-53.5) | 38  (32-45) | 0.0254 |
|  | Min-Max | 24-67 | 33-65 | 34-66 | 31-55 |  |
| PROMIS: Sleep Disturbance | Median (IQR) | 53.8  (48.7-59.8) | 49.1  (44.2-56) | 48.8  (43.3-50.9) | 60.3  (49.5-64) | 0.0638 |
|  | Min-Max | 30.5-70 | 30.5-63.8 | 36.1-58.2 | 42.4-72 |  |
| PROMIS: Sleep-Related Impairment | Median (IQR) | 55.75  (51.1-61.6) | 54.45  (46.1-59.3) | 48.7  (42.1-53.9) | 60.4  (52.5-67.1) | 0.0398 |
|  | Min-Max | 30-79.9 | 30-63.7 | 30-57.9 | 49.2-71.9 |  |
| DSM-5 PTSD Checklist Total Score | Median (IQR) | 20  (8-49) | 2  (0-12) | 1.5  (0-4) | 47  (31-52) | <0.001 |
|  | Min-Max | 0-68 | 0-48 | 0-15 | 25-62 |  |

**Figure 2 NIH Toolbox: Cognition Fluid Composite**

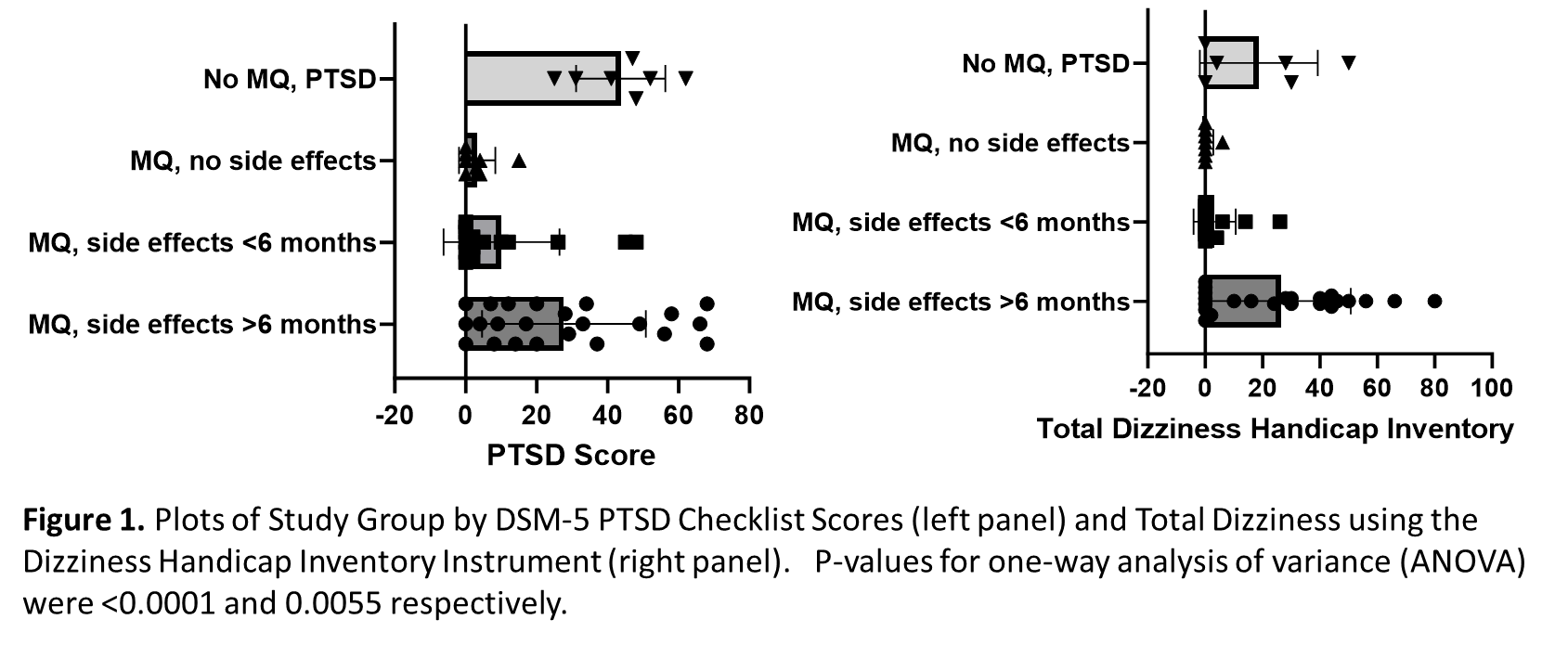

Figure 3: Plots of Study Group by DSM-5 PTSD Checklist Scores (left panel) and Total Dizziness Handicap Inventory (right panel). P-values for one-way analysis of variance (ANOVA) were <0.0001 and 0.0055 respectively.

**Supplementary Tables**

**ORM1RS17650**

| ITEM |  | Homozygous  Reference Alleles | Heterozygous | Homozygous  Alternative Alleles | P-value |
| --- | --- | --- | --- | --- | --- |
| DHIS F19 | No | 10 | 23 | 6 | 0.024 |
|  | Sometimes | 0 | 5 | 4 |  |
|  | Yes | 1 | 0 | 2 |  |

**ORM1RS1126801**

| ITEM |  | Homozygous  Reference Alleles | Heterozygous | Homozygous  Alternative Alleles | P-value |
| --- | --- | --- | --- | --- | --- |
| DHIS P4 | No | 45 | 1 | - | 0.023 |
|  | Sometimes | 2 | 2 | - |  |
|  | Yes | 1 | 0 | - |  |

**MTHFRRS1801133**

| ITEM |  | Homozygous  Reference Alleles | Heterozygous | Homozygous  Alternative Alleles | P-value |
| --- | --- | --- | --- | --- | --- |
| DHIS F3 | No | 21 | 19 | 2 | 0.007 |
|  | Sometimes | 0 | 1 | 3 |  |
|  | Yes | 3 | 2 | 0 |  |
| DHIS P17 | No | 22 | 21 | 2 | 0.009 |
|  | Sometimes | 2 | 1 | 3 |  |
|  | Yes | - | - | - |  |

**MDR1RS1045642**

| ITEM |  | Homozygous  Reference Alleles | Heterozygous | Homozygous  Alternative Alleles | P-value |
| --- | --- | --- | --- | --- | --- |
| DHIS E23 | No | 7 | 24 | 8 | 0.012 |
|  | Sometimes | 0 | 3 | 7 |  |
|  | Yes | 1 | 1 | 0 |  |

**HT2ARS7997012**

| ITEM |  | Homozygous  Reference Alleles | Heterozygous | Homozygous  Alternative Alleles | P-value |
| --- | --- | --- | --- | --- | --- |
| DHIS E23 | No | 2 | 20 | 9 | 0.017 |
|  | Sometimes | 2 | 1 | 6 |  |
|  | Yes | 2 | 3 | 6 |  |

**List of Abbreviations**

| **Abbreviation** | **Term** |
| --- | --- |
| 5HT2A | 5-Hydroxytryptamine (Serotonin) 2A |
| AAG | Alpha_1_-Acid Glycoprotein |
| ADA | Adenosine deaminase |
| ADORA2A | Adenosine 2-α Receptor |
| AEs | Adverse Events |
| BMI | Body Mass Index |
| CAKβ | Cell Adhesion Kinase β |
| CATK | Calcium-Dependent Tyrosine Kinase |
| CNS | Central Nervous System |
| DHI | Dizziness Handicap Inventory Scale |
| DSM-5 PTSD | Diagnostic and Statistical Manual of Mental Disorders, 5^th^ edition—Post-Traumatic Stress Disorder |
| eNOS | Endothelium-Derived Nitric Oxide |
| FAK | Focal Adhesion Kinase |
| FDA | Food and Drug Administration |
| GBM | Glioblastoma Multiforme |
| GVCF | Genomic Variant Call Format |
| GWAS | Genome-Wide Association Studies |
| MAF | Minor Allele Frequency |
| MAP | Mitogen-Activated Protein |
| MDR1 | Human Multidrug Resistance Gene |
| MQ | Mefloquine |
| MTHFR | Methylenetetrahydrofolate Reductase |
| NMDA | N-Methyl D-Aspartate |
| ORM1 | Human Orosomucoid (Alpha_1_-Acid Glycoprotein) |
| PCA | Principal Component Analysis |
| P-gp | P-glycoprotein |
| PROMIS | Patient-Reported Outcomes Measurement Information System |
| PTSD | Post-Traumatic Stress Disorder |
| Pyk2 | Nonreceptor tyrosine kinase |
| RAFTK | Related Adhesion Focal Tyrosine Kinase |
| REDCAP | Research Electronic Data Capture |
| RNA | Ribonucleic Acid |
| RSID | Refence SNP IDs |
| SKAT | Sequence Kernel Association Test for Gene Enrichment |
| SKAT-O | Adaptive Sum of SKAT and Burden Test |
| SNP | Single Nucleotide Polymorphism |
| SNV | Single Nucleotide Variant |
| TBI | Traumatic Brain Injury |
| TOMM | Test of Memory Malingering |
| VCF | Variant Call Format |
| VSSF | Vertigo Symptom Scale—Short Form |
| WGS | Whole Genome Sequencing |
